# Screening for common mental disorders in caregivers of children with severe acute malnutrition in a nutritional rehabilitation centre using the WHO Self-Reporting Questionnaire-20

**DOI:** 10.1101/2024.05.22.24307753

**Authors:** Radhika Mathur, Nisha Kamble, Saba Pathan, Tejal Lakhan, Ayushi Shah, Sanjay Prabhu, Minnie Bodhanwala

## Abstract

A high number of caregivers of children admitted for inpatient treatment of Severe Acute Malnutrition (SAM) at the Nutritional Rehabilitation Centre (NRC), experience psychological distress. It is essential for the caregiver, generally the mother, to feed, hold, comfort, and play with her child as much as possible to promote opportunities for early learning and faster recovery from SAM. Hence, identification of mental health issues among the caregivers is imperative. Using WHO Self-reporting questionnaire (SRQ) 20 score to screen for common mental disorders (CMD) of caregivers of children admitted with SAM. The primary caregiver was interviewed using WHO SRQ-20 scoring. Based on the questions answered YES, the total score was given. A total score of > 8 was considered as a positive screen. These positive-screened caregivers were forwarded to a psychologist. We also compared the CMD prevalence between caregivers of children with primary SAM versus those with secondary SAM. There were a total of 29.1% (n=60) of the caregivers who were screened positive for CMD. The category of CMD most prevalent on screening was somatic issues, followed by depression, anxiety, and cognitive symptoms. Caregivers of children with pre-existing co-morbidity were higher in number than those without one on a positive screen. Maternal mental health is widely neglected, especially in an NRC setting across the country. For mothers of children with severe wasting, underlying mental stressors are significant contributors to the child’s malnourished state. Through this study, we hope to incorporate maternal mental health screening in NRCs for better management of these cases.

## Introduction

The WHO definition of maternal mental health is ‘a state of well-being in which a mother realizes her own abilities, can cope with the normal stresses of life, can work productively and fruitfully and is able to make a contribution to her community’ [1].

Maternal or caregivers’ mental health can significantly affect the nutrition of the child through various mechanisms. Breastfeeding being the initial mainstay for providing adequate nutrition to the child, maternal mental health, particularly stress and anxiety, can impact breastfeeding. Perinatal depression and anxiety are common, with prevalence rates for major and minor depression up to almost 20% during pregnancy and the first 3 months postpartum. In addition, perinatal psychiatric disorders hinder a woman’s functioning and correlate with less-than-optimal development in her offspring [2]. It was also acknowledged that the presence of common mental disorders (CMD) during pregnancy predicts postpartum depression and anxiety disorders, highlighting the underdiagnosis and undertreatment of this condition [3].

Extrapolating perinatal mental health to the mental health of mothers with malnourished children, we can assume similar findings. About 80% of brain growth occurs within the first 2 years of life. A parent, grandparent, or other caring adult plays the most important role in supporting the emotional and cognitive development of children. Children with severe acute malnutrition (SAM) have an elevated risk of mortality and morbidity. There has been a manifold increase in SAM prevalence (7.7%) in several Indian districts as per the NFHS -5 data, which requires urgent policy response [4]. Responsive parenting and sensory stimulation play a key role in recovery by reducing the risk of developmental and emotional problems in children with SAM, which is also noted in SAM management guidelines [5].

However, there is data showing a high number of caregivers of children admitted for inpatient treatment of SAM experiencing psychological distress, depressive symptoms, and suicidal behaviour [6]. When mothers are involved in the overall care of their child at the hospital, they learn how to continue caring for their children at home. This study aimed to assess the burden of caregiver mental health status among caregivers who are admitted to the nutritional rehabilitation department with their children who are severely wasted.

## Materials and Methods

This study is a cross-sectional observational research study conducted at a tertiary care children hospital Nutritional Rehabilitation Centre (NRC) from 25^th^ July 2023 to 25^th^ March 2024. We included the primary caregivers of all children, who were majorly mothers. An approval from the Institutional Ethics Committee–Human Research was obtained before the commencement of the study. We excluded the cases whose primary caregiver could not be present with the child during the assessment. As per the standard of care, all children admitted to the NRC were treated based on the WHO guidelines adapted by the Ministry of Health and Family Welfare, Government of India [5] for the treatment of SAM children. SAM is defined as children 6 weeks to 6 months with weight-for-age (WFA) < -2SD or weight-for-height (WFH) < -2 SD, or severe visible wasting or nutritional edema; 6 months to 59 months WFH <-3SD, visible severe wasting or nutritional edema [7].

At the time of admission, the primary caregiver, the attendant who stayed with the admitted child, was interviewed using the WHO Self Reporting Questionnaire (SRQ) 20 scoring (8) screening tool by the NRC staff after sharing the study information sheet and taking consent for the same. Based on the questions answered YES, the total score was given, wherein 1 point indicate a YES and 0 was NO. A total score of more than 8 was considered to be a positive screen. The answers were given based on the last 1-month experience of the caregiver. Only screening for CMD was carried out and no diagnosis was made. Based on the specific questions answered YES, the issues were divided into cognitive symptoms, anxiety, depression, and somatoform issues based on the user guidebook of the screening tool [8], and this was not disclosed to the person screened.

These positive-screened caregivers were then referred to the Clinical Psychologist for an in-depth screening for anxiety and depression disorders based on the Diagnostic and Statistical Manual of Mental Disorders – 5^th^ Edition (DSM 5) [9]. A self-designed, psychological proforma was used for identifying the basic antenatal and postnatal concerns and significant stressors based on the biopsychosocial model. Mental Status Examination (MSE) was employed for each caregiver [10]. The stressors were categorized into 3 categories, i.e., child-related, family-related, and social (lack of social/financial support). The severity of stressors was marked on a Likert scale ranging from 0–4 (0-not applicable, 1-mild, 2-moderate, 3-severe, 4-very severe). Suicide ideations were marked on a binary scale of 0 or 1 (0-no, 1-yes). Based on the outcome of the interview, the caregivers were provided counselling therapy and offered the option to continue the therapy. Those caregivers identified to have very severe ratings were then referred to the psychiatrist for necessary pharmacological treatment.

Statistical analysis: Data was analysed using SPSS (Version 21.0) software. Categorical data was summarized as percentages, whereas continuous data was summarized using mean and standard deviation (SD). Mean scores between different subgroups were compared using the unpaired t-test. The association between two categorical variables was determined using the chi-square test. Multivariate analysis was performed using the linear regression model as the dependent variable is continuous in nature.

### Ethics Statement

The study was conducted after institutional ethics approval and after obtaining written informed consent from the study participants.

## Results

A total of 206 caregivers were screened, of which 29.1% (n=60) had a score of greater than 8, i.e., screened positive for CMD and all of them were mothers of the child admitted. The category of CMD which was most prevalent on screening among the ones who were positive were somatic (Mean: 4.35±1.58; Median: 4) followed by depression (Mean: 4.30±1.63; Median: 4), anxiety (Mean: 2.18±0.81; Median: 2), and cognition (Mean: 1.63±0.96; Median: 2) based on the WHO SRQ-20 screening tool. Caregivers of children with pre-existing co-morbidity (neurologically impaired, congenital heart disease, structural co-morbidities, etc.) were higher in number than ones without co-morbidity on a positive screen. Out of positive 60 screened, 26 had children less than 6 months of age. Out of 60 mothers who were evaluated and screened positive for CMD using WHO SRQ-20, 33 caregivers were interviewed further to understand the associated factors of CMD. These 33 mothers were selected as it was difficult to follow up with all the mothers. Among the mothers screened positive on the WHO SRQ-20 screening tool, 87.9% (29/33) were from a low socioeconomic stratum, 18.2% (6/33) were educated under 10^th^ standard, 16% (16/33) up to 10^th^ standard, 18.2% (2/33) up to 12^th^ Standard. Additionally, 21.2% (7/33) had more than three children and 12.1% (4/33) had miscarriages in the past. On interviewing the mothers, the predominant cause of the mental health issues on the Likert scale included child-related (75.8%-severe), family-related (24.2%-mild) and social causes like lack of family support, poverty (36.4%-severe). Among the positively screened, 51.3% and 30.3% had depression and anxiety features, respectively, based on the DSM 5.

The Table 1 represents the results of a study investigating various factors related to the need for psychiatric evaluation among mothers who screened positive.

**Table 1:** Risk factors in the caregiver requiring psychiatric evaluation.

| **Psychiatric evaluation needed** |  |  |  |  |
| --- | --- | --- | --- | --- |
| **Maternal Age at Delivery** | No (%) | Yes (%) | N | p-value |
| <=20 yrs | 75.0 | 25.0 | 4 | .845 |
| 21-25 yrs | 69.2 | 30.8 | 13 |  |
| 26-30 yrs | 54.5 | 45.5 | 11 |  |
| >30 yrs | 60.0 | 40.0 | 5 |  |
| **Birth Order** |  |  |  |  |
| 1 | 86.7 | 13.3 | 15 | .070* |
| 2 | 54.5 | 45.5 | 11 |  |
| 3 | 50.0 | 50.0 | 2 |  |
| 4 | 33.3 | 66.7 | 3 |  |
| 5 | 0.0 | 100.0 | 2 |  |
| **Suicide ideations** |  |  |  |  |
| Absent | 73.1 | 26.9 | 26 | .030** |
| Present | 28.6 | 71.4 | 7 |  |
| **Support** |  |  |  |  |
| No | 25.0 | 75.0 | 4 | .087* |
| Yes | 69.0 | 31.0 | 29 |  |
| **Socio-economic Status** |  |  |  |  |
| Low | 65.5 | 34.5 | 29 | .545 |
| Middle | 50.0 | 50.0 | 4 |  |
| **Education** |  |  |  |  |
| 5<sup>th</sup> std | 0.0 | 100.0 | 3 | .094* |
| 8<sup>th</sup> std | 33.3 | 66.7 | 3 |  |
| 10<sup>th</sup> std | 75.0 | 25.0 | 16 |  |
| 12<sup>th</sup> std | 66.7 | 33.3 | 6 |  |
| 15<sup>th</sup> std | 80.0 | 20.0 | 5 |  |
| **Miscarriage** |  |  |  |  |
| None | 65.5 | 34.5 | 29 | .405 |
| 1 | 66.7 | 33.3 | 3 |  |
| 2 | 0.0 | 100.0 | 1 |  |
| **Child Death** |  |  |  |  |
| None | 66.7 | 33.3 | 27 | .566 |
| 1 | 66.7 | 33.3 | 3 |  |
| 2 | 50.0 | 50.0 | 2 |  |
| 3 | 0.0 | 100.0 | 1 |  |
| **Antenatal care** |  |  |  |  |
| No | 62.5 | 37.5 | 32 | .443 |
| Yes | 100.0 | 0.0 | 1 |  |
| **Anxiety** |  |  |  |  |
| No | 100.0 | 0.0 | 3 | .170 |
| Yes | 60.0 | 40.0 | 30 |  |
| Note: p-values are from Chi-square test |  |  |  |  |
| *p-value significant at 90% C.I. |  |  |  |  |
| **p-value significant at 95% C.I. |  |  |  |  |

It can be observed that individuals with higher birth orders may be more likely to require psychiatric evaluation. Also, there is a significant association between suicide ideations and the need for psychiatric evaluation. Individuals reporting suicide ideations are much more likely to require evaluation. The table also indicates that individuals lacking support and poor education may be more likely to need psychiatric evaluation, and the result is statistically significant. However, many other factors examined in the study do not appear to have significant associations with the need for psychiatric evaluation.

## Discussion

In our study, we found a prevalence of 29.1% of caregivers who were screened positive for CMD. A study showed that maternal CMD (MCMD) prevalence was as high as 40.7% in out-patient malnutrition clinics in North-western Nigeria [11]. A case-control study including 294 children aged between 0 and 5 years was conducted in Northeast Brazil using the SRQ-20 screening tool on caregivers of children diagnosed as moderately or severely malnourished. MCMD doubled the risk of moderate or severe malnutrition in children (OR=2.04; 95% CI: 1.10–3.78), thereby proving that maternal caregiving can be an important factor mediating the relationship between maternal mental health and child malnutrition. CMDs may impair the mother’s ability to perform her role, particularly in deprived environments [12]. In a study conducted by Samuel Scott et al, it was found that CMD, which was defined as ≥ 8 positive SRQ responses, was reported by 262 women (16%) [13].

We found the category of CMD most prevalent on screening were somatic issues followed by depression, anxiety, and cognitive symptoms, in that order. Caregivers of children with pre-existing co-morbidity (neurologically impaired, congenital heart disease, structural co-morbidities, etc) were higher in number than those without co-morbidities on a positive screen. In a study by Ejaz et al, anxiety and depression are prevalent psychiatric conditions in our society, as indicated by significantly elevated Hospital Anxiety and Depression Scale (HADS) scores among mothers. Predictors of higher HADS scores in mothers of malnourished children include advancing maternal age, low birth weight of the child, larger family size, and low income [14]. A systematic review of studies by Stewert et al indicated a robust correlation between maternal depressive symptoms, assessed using tools like the WHO SRQ-20, and infant undernutrition, particularly pronounced within low socio-economic communities where women encounter heightened adversities and experience lower levels of empowerment [15]. In a similar study by Rahman A. et al, infants exposed to maternal mental distress, as determined by the WHO SRQ-20, face an elevated risk of undernutrition [16]. A multinational study by Nguyen et al also showed that MCMD, affecting nearly half of women in Bangladesh and one-third in Vietnam, emerged as a significant factor contributing to child stunting and underweight, respectively. Although CMD affected 39% of women in Ethiopia, no similar association with child undernutrition was observed. However, maternal CMD showed a robust association with childhood illnesses across all three countries [17]. A study by Miranda et al indicated that the converse, i.e., mothers of children with protein energy malnutrition (PEM) showed a higher rate of mental disturbances than mothers of eutrophic children [18].

As risk factors, we observed that individuals with higher birth orders may be more likely to require psychiatric evaluation. Also, there is a significant association between suicide ideations and the need for psychiatric evaluation. Individuals reporting suicide ideations are much more likely to require evaluation. In a study by Alao et al, antenatal care at primary healthcare facilities (adjusted odds ratio [aOR: 13], primary education [aOR: 3.255] living in the south-southern region of the country [aOR: 2.207], poor breastfeeding support [aOR: 1.467], polygamous family settings [aOR: 2.182], and a previous history of mental health disorders [aOR: 4.684] were significantly associated with CMD. In contrast, those from the middle and lower socioeconomic classes were less likely to develop CMD, with [aOR: 0.532] and [aOR: 0.493], respectively [19]. Additionally, several factors can contribute to maternal mental health issues when caring for malnourished children, including emotional stress due to their child with co-morbid conditions like congenital heart disease, cerebral palsy, chronic lung disease, anatomical defects, etc., can be emotionally distressing for mothers. This was also observed in our study for mothers whose children had secondary SAM. They may experience feelings of guilt, helplessness, and worry about their child’s health and future. Mothers may experience stress and anxiety related to financial difficulties, including concerns about affording nutritious food, medical treatment, and other basic needs for their child.

Malnutrition can affect the parent-child relationship, as mothers may feel distressed or inadequate if they perceive themselves as failing to meet their child’s nutritional needs. Maternal health can also be directly affected by the stress and strain of caring for a malnourished child. Addressing maternal mental health is essential for the well-being of both mothers and their malnourished children. Providing mothers with emotional support, access to mental health services, and interventions to alleviate financial strain and social isolation can help mitigate the impact of maternal mental health issues on caregiving and child health outcomes.

Moving forward, it is imperative that maternal or caregiver mental health be looked at closely especially in a nutritional rehabilitation set up. A study by Rahman A et al showed that interventions administered by trained non-specialist health and community workers demonstrated greater efficacy than standard care for both mothers and children. Furthermore, where evaluated, benefits to the child encompassed enhanced mother-infant interaction, improved cognitive development and growth, decreased incidences of diarrheal episodes, and higher rates of immunization [20]. Maternal mental health services must be integrated into the facility-based management, i.e., NRC management of acute malnutrition programs to provide more holistic care and possibly improve long-term outcomes after discharge from these programs.

## Conclusion

CMD among caregivers affects child nutrition and developmental status and contributes to delayed resolution from malnourishment. It has been seen that the prevalence of CMDs in caregivers of SAM children is high in India. The WHO SRQ-20 validated screening tool could be considered as a standardised tool to be used in all NRCs. A forward plan must be established for the mothers who have been screened positive for CMDs from NRCs. Importance of screening all mothers with children admitted in NRCs for CMD should be emphasized.

## Data Availability

Authors confirm and abide by the PLOS Data Policy. Data pertaining to this study can be accessed as per policy.

